# Spatial clustering of undervaccination amplifies local measles transmission but limits geographic spread: South Carolina, 2025–2026

**DOI:** 10.64898/2026.09.21.26363346

**Authors:** Aakash Pandey, Amanda M Bleichrodt, Emily A Serman, Shakhawat H. Tanim, Ana I. Bento, Lior Rennert

**Affiliations:** Department of Public Health Sciences, Clemson University, SC, USA; Department of Public and Ecosystem Health, Cornell University, NY, USA

## Abstract

The 2025 measles outbreak in South Carolina, the largest in the United States in over two decades, produced 997 confirmed cases, predominantly among school-aged children in an under-vaccinated community in Spartanburg County. To support the South Carolina Department of Public Health (DPH) measles response, we developed a stochastic agent-based model to forecast outbreak trajectory and evaluate intervention strategies in real time, integrating transmission across three spatial scales and grounded in geocoded households, school enrollment records, and census-derived demographic data. The model predicted a median of 1,020 total infections (90% prediction interval: 840–1,273). Counterfactual simulations without active contact tracing and quarantine projected a median of 2,432 infections (90% prediction interval: 1,867–3,185), approximately 138% more than the model-predicted baseline (1,020 cases). Model fidelity required a highly localized between-school contact structure; broader contact assumptions substantially overpredicted both case counts and geographic spread. Simulated regional introductions confirmed that outbreaks arose only near connected clusters of undervaccination, while well-vaccinated areas remained contained. Model versions deployed prospectively at weeks 9, 17, and 34 of the response informed resource allocation decisions and provided operational evidence for the effectiveness of contact tracing and quarantine strategies. Critically, the model reproduced geographic spread, number of affected schools, and breakthrough infection counts not used in fitting, validating its structural fidelity and establishing a replicable framework for jurisdictions maintaining school-enrollment and vaccination records.

## Introduction

Measles, eliminated in the United States in 2000, has resurged as international importations of the virus take hold in localized, spatially-concentrated populations with undervaccination (1, 2). The 2025 South Carolina outbreak produced 997 confirmed cases predominantly among school-aged children in an under-vaccinated community in Spartanburg County, making it one of the largest reported single-jurisdiction outbreaks in the United States in more than three decades (3, 4). Containing such outbreaks benefits from real-time quantitative tools capable of forecasting case burden, evaluating intervention scenarios, and estimating impact (5), yet to our knowledge, the prospective deployment of spatially explicit agent-based models grounded in local surveillance and demographic data within an active public health emergency response has rarely been reported.

Here we report a stochastic agent-based model iteratively developed and prospectively deployed within the DPH Measles Incident Management Team and show that its retrospective calibration to a single contact-network parameter reproduced multiple outbreak features not used in fitting, enabling quantitative evaluation of intervention impact.

## Results

### Model validation against observed outbreak

A localized between-school contact structure was necessary to reproduce observed outbreak dynamics: predicted size was highly sensitive to the assumed driving-time threshold, with a best-fit value of 19 minutes, between the first quartile (16 min) and median (22 min) of inter-school driving times in Spartanburg County (Fig. 1A). Lower thresholds underpredicted and higher thresholds substantially overpredicted case counts and geographic spread. This threshold predicted a median of 1,020 infections (90% prediction interval: 840–1,273), a median of 30 affected schools (90% prediction interval: 20–41) compared to 33 observed, and a median of 11 breakthrough infections (90% prediction interval: 4–20) compared to 20 observed among two-dose MMR recipients. Geographic spread beyond Spartanburg County was a stochastic outcome, occurring in 42% of simulations for Greenville County and fewer than 5% of simulations for all other counties in the study region (Fig. 1B). In simulations where spread reached Greenville County, the model predicted a median of 34 cases compared to 36 observed, while Spartanburg County accounted for a median of 1,010 predicted cases compared to 940 observed (Fig. 1C and 1D).

**Figure 1.**
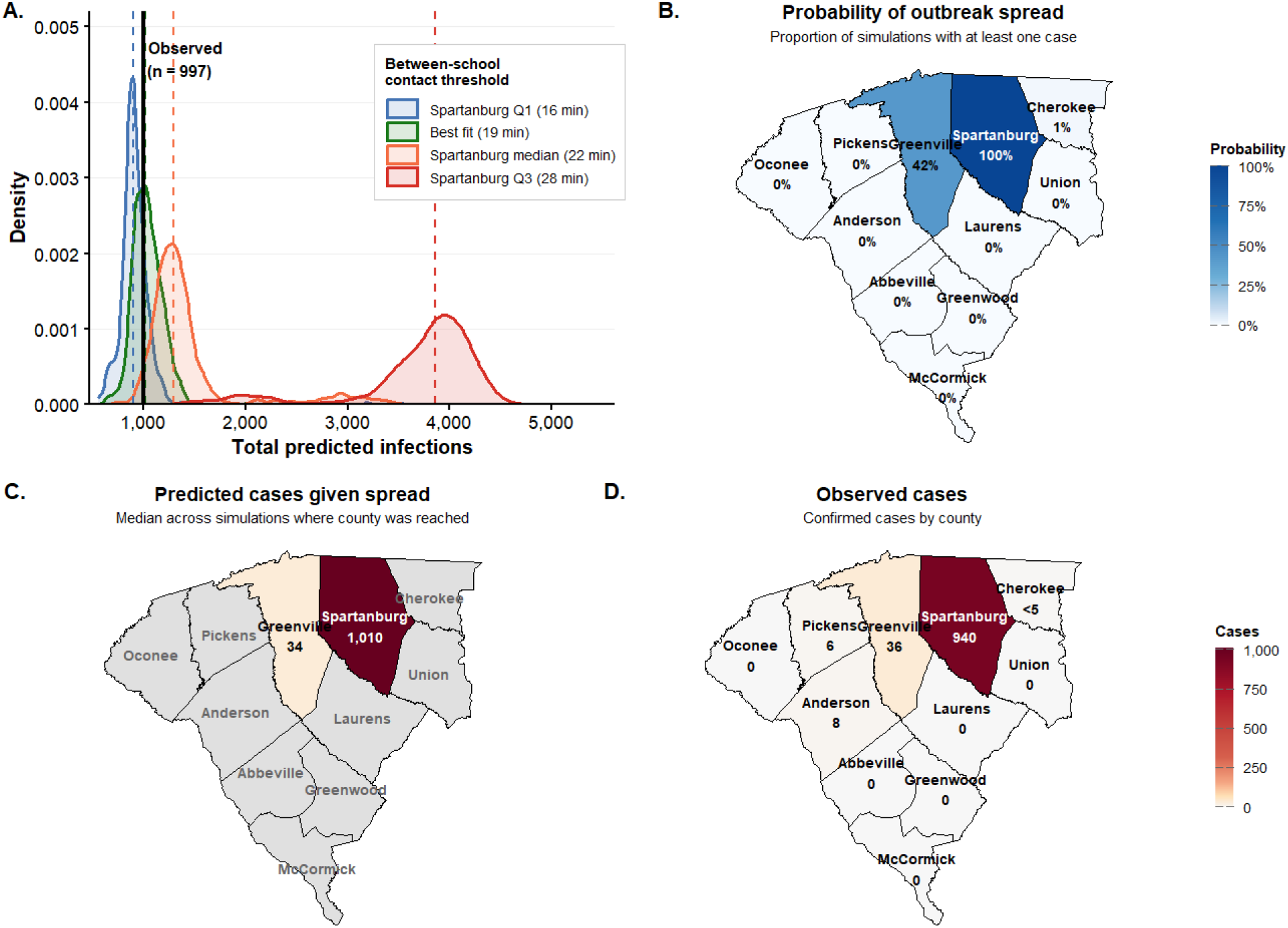
Simulated measles outbreak dynamics and spatial spread in Upstate South Carolina. (A) Predicted final outbreak size distributions across four between-school contact thresholds, with the observed case count (n = 997) shown as a vertical reference line; dashed lines indicate scenario medians. (B) Probability of outbreak spread by county under the best-fit model (19-minute threshold), defined as the proportion of 1,000 simulations producing at least one case. (C) Median predicted case count by county conditional on outbreak spread, shown only for counties where spread probability exceeded 5%; grey counties fell below this threshold. (D) Confirmed case counts by county.

Simulated introductions into an under-vaccinated cluster (Anderson County) versus a higher-coverage area (Oconee County) confirmed that introduction location governs large-outbreak risk. Both scenarios produced small outbreaks in most simulations but differed markedly in the risk of a large outbreak: introduction into the Anderson County hotspot exceeded 100 cases in 12% of simulations (median 9 cases, 90% prediction interval: 5–1,810), whereas introduction into the Oconee County non-hotspot exceeded 100 cases in only 0.3% of simulations (median 11 cases, 90% prediction interval: 6–49).

### Impact of public health response

To quantify the contribution of the organized public health response to outbreak containment, we performed counterfactual simulations in which case isolation occurred only at rash onset with no contact tracing or quarantine. Under this scenario, the model predicted a median of 2,432 infections (90% prediction interval: 1,867–3,185), compared to 1,020 under the default response (Fig. 2A). Reducing the isolation delay from 4 to 2 days, reflecting the improvement in detection speed observed during the later phase of the outbreak, produced only a modest change in median outbreak size (Fig. 2A). This limited effect likely reflects two features of the response: high efficacy contact tracing already identified and quarantined most exposed contacts regardless of isolation timing, and isolated cases continued to transmit within their households, a pathway unaffected by faster detection. Outbreak size was substantially more sensitive to quarantine efficacy: at the lowest efficacy evaluated (*K* = 0.25), the model predicted a median of 1,542 infections (90% prediction interval: 1,167–1,890), with predictions declining monotonically as efficacy increased to *K* = 0.90 (Fig. 2B).

**Figure 2.**
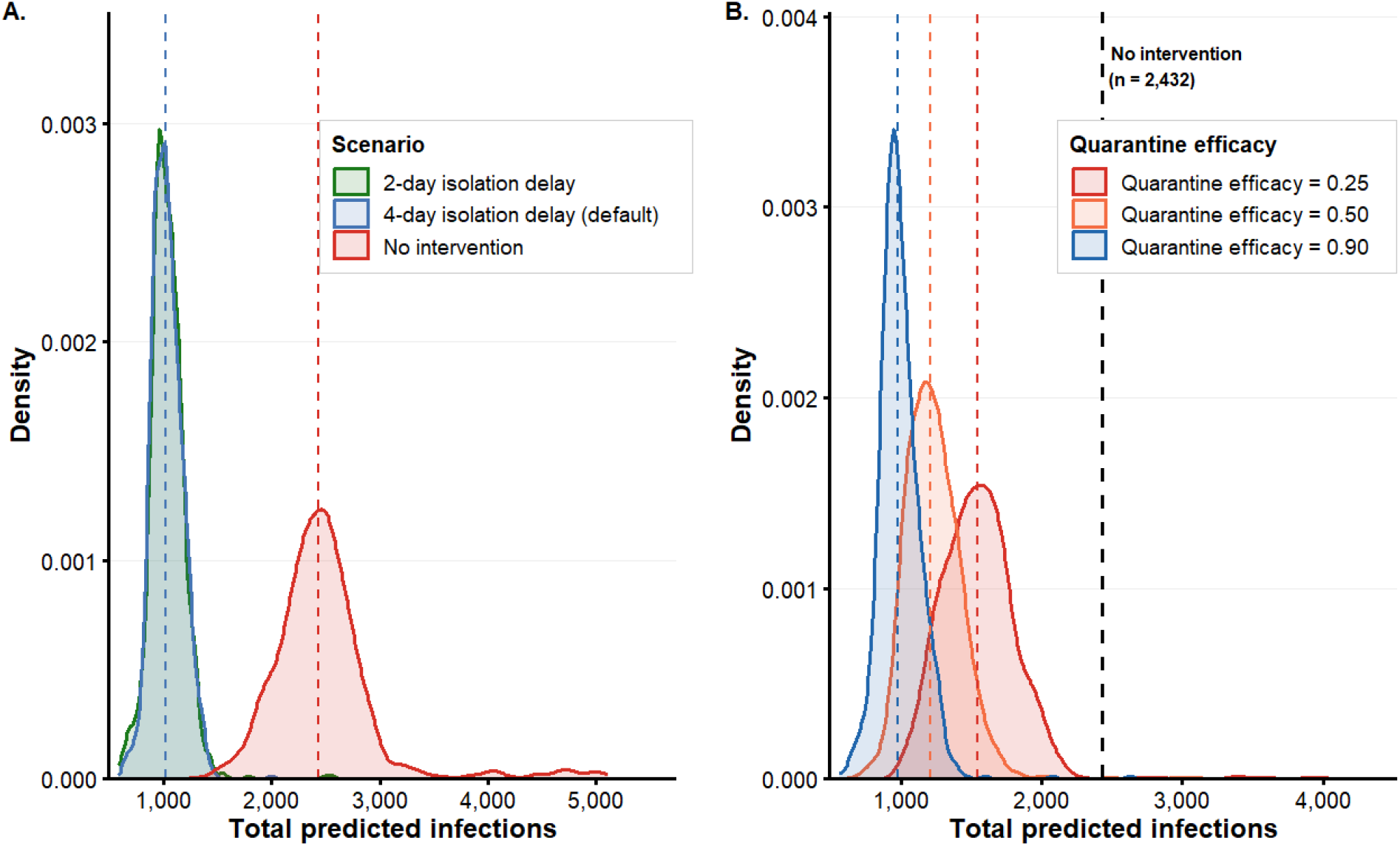
Impact of public health intervention parameters on predicted outbreak size. (A) Predicted final outbreak size distributions under three isolation delay scenarios: 2-day delay, 4-day delay (default), and no intervention; dashed lines indicate scenario medians. (B) Predicted final outbreak size distributions under three quarantine efficacy levels; dashed black line indicates the no-intervention median for reference.

## Discussion

The close agreement between model predictions and observed outcomes, in both total case burden and geographic concentration, reflects a key structural feature of this outbreak: transmission was driven primarily by fine-scale spatial clustering of undervaccination that aggregate county-level coverage statistics fundamentally obscure (6, 7). At the outbreak’s onset, Spartanburg County’s average school vaccination coverage was approximately 88%, yet individual school coverage ranged from 21% to 100% (8), a heterogeneity that county-level averages cannot capture but that determines where transmission can be sustained. That broader contact assumptions failed to reproduce observed dynamics independently corroborates this mechanism (7) and confirms that spatially resolved models are most valuable precisely where aggregate surveillance data are least informative. The relative insensitivity of outbreak size to isolation delay compared to quarantine efficacy further suggests that contact tracing capacity, rather than speed of individual case detection, is the primary operational lever for containment of school-based measles outbreaks, with direct implications for resource allocation during future responses (9). This clustering simultaneously amplified within-community transmission while limiting spread beyond it, producing outbreaks that are intense but spatially bounded and amenable to geographically targeted response.

Several limitations merit consideration. The model represents school and household transmission only; community settings, including places of worship and recreational facilities that contributed to the spread, are not explicitly modeled. Two omissions warrant particular attention because they bias the model in opposing directions. A mid-outbreak behavioral change, likely associated with holiday gatherings, produced a case spike the model did not capture, tending to underestimate transmission. Conversely, the model does not represent the reactive vaccination campaign mounted during the response, which administered 3,788 additional MMR doses in Spartanburg County and 14,745 across Upstate South Carolina over the preceding year (increases of 94% and 82% respectively), tending to overestimate transmission by omitting the resulting reduction in susceptibility. These opposing biases may partially offset. Confidence in model fidelity therefore rests principally on agreement across the independent validation targets: geographic distribution, school count, and breakthrough infections, rather than the aggregate total alone.

As domestically acquired measles cases continue to rise, public health agencies require forecasting tools that can be deployed within the time and data constraints of an active response. This model was developed and deployed in successive versions during the outbreak: a school-level outbreak risk tool at week 9, an inter-school transmission model at week 17, and the full household-integrated model at week 34, each shared with response leadership as it became available. At each deployment, model forecasts bracketed the outbreak’s eventual 997 confirmed cases: the inter-school model (week 17) predicted 1,297 median cases (90% PI: 797–1,685), and the full household-integrated model (week 34) predicted 1,003 median cases (90% PI: 996–1,275). Model projections helped leadership assess whether anticipated outbreak scenarios fell within a manageable planning range, informing decisions to redeploy staff to the Upstate region and reallocate routine work toward outbreak response, and provided evidence that contact tracing, isolation, and quarantine were effective. A school-level forecasting tool derived from the model supported local planning by administrators across the state. Although calibrated to a single outbreak, the model’s reproduction of confinement across independently identified undervaccinated and well-vaccinated areas suggests its structure captures generalizable features of spatially clustered measles transmission. This work demonstrates that an agent-based model grounded in school enrollment records, geocoded census populations, and routine surveillance data can be developed and iterated within an active incident management structure to deliver operational forecasts, evaluate intervention scenarios, and quantify response impact in real time, offering an approach adaptable to future outbreaks in similar settings.

## Methods

A full description of the model is provided in SI Methods. The simulation model code is publicly available at GitHub (https://github.com/DMA-PRIME/measles-household-school-cluster).

## Data Availability

The simulation model code is publicly available at GitHub (https://github.com/DMA-PRIME/measles-household-school-cluster).

## Acknowledgments

South Carolina Department of Public Health 2025 Measles Incident Management Team; Dr. Marco E. Tori, Dr. Linda Bell; Dr. Edward Simmer.

## References

1. CDC, Vaccination coverasssge and exemptions among kindergartners. SchoolVaxView (2025). Available at: https://www.cdc.gov/schoolvaxview/data/index.html [Accessed 20 May 2026].

2. M. Fattah, et al., Trends in county-level childhood vaccination exemptions in the US. JAMA 335, 546–549 (2026).

3. SCDPH, DPH announces end to measles outbreak in Upstate at 997 cases. Available at: https://dph.sc.gov/news/dph-announces-end-measles-outbreak-upstate-997-cases [Accessed 20 May 2026].

4. CDC, Measles cases and outbreaks. Measles (Rubeola) (2026). Available at: https://www.cdc.gov/measles/data-research/index.html [Accessed 20 May 2026].

5. N. B. Masters, Real-time use of a dynamic model to measure the impact of public health interventions on measles outbreak size and duration — Chicago, Illinois, 2024. MMWR Morb Mortal Wkly Rep 73 (2024).

6. N. B. Masters, et al., Fine-scale spatial clustering of measles nonvaccination that increases outbreak potential is obscured by aggregated reporting data. Proceedings of the National Academy of Sciences 117, 28506–28514 (2020).

7. E. A. Serman, B. Witrick, L. Rennert, Clusters of concern — Spatial link between childhood undervaccination and measles outbreaks in South Carolina. New England Journal of Medicine 0.

8. SC Immunization Data | South Carolina Department of Public Health. Available at: https://dph.sc.gov/public/vaccinations/sc-immunization-data [Accessed 21 May 2026].

9. W. T. A. Enanoria, et al., The effect of contact investigations and public health interventions in the control and prevention of measles transmission: A simulation study. PLOS ONE 11, e0167160 (2016).

