## Supplemental Methods for "Spatial clustering of undervaccination amplifies local measles transmission but limits geographic spread: South Carolina, 2025–2026"

**Model Overview**

We developed a discrete-time stochastic agent-based network model to simulate measles transmission among school-aged children and their household contacts across 11 counties in Upstate South Carolina, with the dual purpose of generating prospective outbreak forecasts and evaluating counterfactual intervention scenarios. The model represents two agent types: school agents, comprising individual students aged 5-18 each assigned to a specific school and geocoded household, and household agents, comprising non-school-age members of households containing at least one enrolled student, including adults and young children. Both agent types progress through the same compartmental structure: susceptible (S), vaccinated (V), latent-exposed (E), prodromal (P), rash ($R_{a}$), isolated (Iso), and recovered (R), with quarantine states (QS, QE) assigned to unvaccinated susceptible and latent contacts of confirmed cases, respectively. School agents participate in within-school, between-school, and household transmission; household agents participate in household transmission only and may seed infections back into the school network via co-resident students. Vaccination is implemented as an all-or-nothing efficacy model with household-correlated uptake, whereby all children within a household share the same vaccination outcome, thereby capturing family-level clustering of susceptibility. Case isolation and contact quarantine are represented explicitly and parameterized from outbreak response records.

**Population Structure**

**School populations**

School-level data, including enrollment counts, reported vaccination coverage, and grade ranges, were obtained from South Carolina Department of Public Health surveillance records (1). Each school's student population was represented as individual agents assigned to classrooms of fixed size $c=25$ students, with ages sampled uniformly from the grade-appropriate range derived from each school's reported minimum and maximum grade. Each student agent carries individual-level attributes including disease compartment state, vaccination status, vaccine efficacy realization (the Bernoulli draw determining whether vaccination confers full protection under the all-or-nothing model), time-in-state counter, infection source label, and household identifier, enabling direct tracking of transmission chains, breakthrough infections, and household-school linkages.

**Synthetic household population**

Household structure was derived from the RTI International synthetic population for South Carolina (2), which provides individual-level records including person identifiers, household identifiers, age, and geocoded household location. School-age children (ages 5–18) in the synthetic population were matched to schools using a two-criterion algorithm. First, each child's grade was estimated as

$$g_{i}=\text{age}_{i}-5$$

and a school was considered grade-appropriate if its reported grade range satisfied $g_{\min}\leq g_{i}+2$and $g_{\max}\geq g_{i}-2$, permitting a tolerance of $\pm2$grades. Second, among all grade-appropriate candidate schools, the nearest was selected by Haversine great-circle distance; children whose grade matched no available school were assigned to the nearest school of any grade. Prior to assignment, the synthetic population was restricted to households within approximately 30 km of study-region schools. This buffer was chosen to exceed plausible home-to-school commuting distances in the Upstate region of South Carolina, where Spartanburg County spans roughly 2,100 km² across seven school districts (mean district radius approximately 10 km), ensuring the eligible population captured realistic school catchments while excluding distant households unlikely to attend study-region schools. Because assignment selected the nearest grade-appropriate school without a per-child distance cutoff, no child within this buffer remained unassigned; only households outside the buffer were excluded.

A subsequent capacity-balancing step identified over-enrolled schools, defined as those exceeding 120% of reported enrollment, and reassigned their most distal excess students to the nearest grade-appropriate alternative until enrollment approached reported levels. After school assignment, all household members residing with at least one school-age child were included in the household transmission population, comprising adults and non-school-age children who participate in within-household transmission but are not enrolled in the school contact network.

Households containing children assigned to different schools create cross-school transmission pathways: a child infected at school may transmit to a sibling at home, who then seeds a distinct school. These sibling-mediated inter-school linkages are an explicit structural feature of the model and represent a mechanistic pathway for geographic spread beyond the index school.

**School Contact Network**

Between-school transmission was mediated by a driving-time-weighted contact network constructed from a precomputed matrix of inter-school driving times for all schools in the study region. An undirected weighted graph was constructed over all schools, where an edge was placed between schools $i$and $j$ if the driving time $t_{ij}$satisfied $t_{ij}\leq d_{\max}$. Edge weights were assigned using an exponential decay function:

$$w_{ij}=\text{ }\text{exp(-}\frac{t_{ij}}{d_{\max}/2}\text{)}$$

capturing the empirical pattern that inter-school social contact rates decline steeply with travel time. The adjacency matrix was symmetrized by taking the maximum of $w_{ij}$and $w_{ji}$.

The threshold $d_{\max}$ governs the geographic scale of between-school contacts and was evaluated across four candidate values anchored to the empirical distribution of inter-school driving times within Spartanburg County: the first quartile ($d_{\max}=16$ min), an intermediate value between the first quartile and median ($d_{\max}=19$ min), the median ($d_{\max}=22$ min), and the third quartile ($d_{\max}=28$ min). Thresholds were drawn from the Spartanburg County distribution rather than the broader Upstate regional distribution, reflecting that transmission-relevant school contacts were confined to the affected community rather than the full study region.

**Disease Natural History**

The model implements a modified SEIR framework with explicit prodromal and rash stages reflecting the natural history of measles. Susceptible individuals progress through the following compartments upon infection: Susceptible (S), Latent-Exposed (E), Prodromal (P), Rash ($R_{a}$), and Recovered (R). Detected cases transition to Isolated (Iso) and are removed from within-school and between-school transmission; however, isolated students remain present in their household and continue to contribute to household transmission during the isolation period. Unvaccinated contacts of confirmed cases enter quarantine states (QS, QE) corresponding to their disease status at the time of quarantine.

Latent period duration $\tau_{E}$was drawn independently for each newly-exposed individual from an Erlang distribution with mean $\mu_{E}=10$ days and shape parameter $k_{E}=8$. Total infectious period duration $\tau_{I}$ was drawn from an Erlang distribution with mean $\mu_{I}=8$ days and shape $k_{I}=8$. The prodromal stage was fixed at $\tau_{P}=4$days, after which the individual entered the rash stage for the residual infectious period $\tau_{Ra}=\max(1,\text{ }\tau_{I}-\tau_{P})$days.

**Transmission Model**

*Within-school transmission*

Within each school, transmission was modeled at two spatial scales: within-classroom and between-classroom. On each simulation day, each non-isolated, non-quarantined infectious individual in state P or $R_{a}$ sampled contacts from the full pool of present students (all non-isolated, non-quarantined individuals regardless of disease state) using Poisson-distributed contact rates $c_{\text{within}}=6 \mathrm{day}^{-1}$ for same-classroom contacts and $c_{\text{between}}=3 \mathrm{day}^{-1}$ for contacts in other classrooms within the same school. Transmission to a susceptible contact occurred with probability $p_{\text{within}}=0.15$ (same classroom) or $p_{\text{between}}=0.10$ (different classroom). Prodromal and rash stages were assigned equal infectiousness. Vaccinated individuals who experienced primary vaccine failure transmitted at reduced infectiousness (reduction factor $\eta=0.80$). A rolling contact history of width 7 days was maintained per school to enable targeted quarantine of contacts upon case isolation.

*Between-school transmission*

On each simulation day, each non-isolated infectious individual at a source school generated inter-school contacts at a base rate $c_{\text{school}}=0.15 \mathrm{day}^{-1}$, scaled by the network edge weight $w_{\mathrm{ij}}$ for each connected school. Contacts were sampled from the full present population of each connected school, and transmission occurred at per-contact probability $p_{\text{between}}=0.10$. The same infectiousness modifiers for vaccine failure applied as in within-school transmission. The number of realized between-school contacts varied stochastically across simulations as a function of the Poisson-distributed contact process and the network structure, contributing to variability in outbreak size and geographic spread across simulation replicates.

*Household transmission*

On each simulation day, each infectious household member (student or non-student) in states P or $R_{a}$ transmitted to each susceptible co-resident with daily probability $p_{\text{hh}}=0.25$. Isolated students remained present in their household and continued to contribute to household transmission during the isolation period, reflecting the practical reality that home isolation concentrates infectious individuals with susceptible household contacts. Non-student household members who became infected progressed through the same compartmental sequence and could transmit back to susceptible household members, including students, creating a bidirectional school-household transmission pathway.

*Parameter justification and implied R_0_*

Contact rates and per-contact transmission probabilities were parameterized to be consistent with empirically derived contact patterns for school-aged populations. Daily within-school contacts ($c_{\text{within}}=6$, $c_{\text{between}}=3$) were derived by decomposing the mean daily contact rate reported for the 10-19 age group (12.7 contacts per day) from the Midwest Analytics and Disease Modeling Center autumn 2025 social contact survey (3), subtracting home (1.7) and community (1.6) contacts, yielding approximately 9.4 school-attributable contacts per student per day, consistent with the model total of 9. Per-contact transmission probabilities were calibrated such that the implied basic reproduction number in a fully susceptible unvaccinated population is roughly 12 to 14 under typical contact and household size assumptions, consistent with published estimates for measles transmission in school-aged populations (12 to 18) (4).

This calibrated value was constrained by the requirement that the model reproduce the observed outbreak rather than selected a priori. The between-school contact threshold analysis reported above functions as a partial sensitivity analysis over effective transmissibility: widening the threshold increases between-school mixing and effective reproduction, and thresholds implying higher transmissibility substantially overpredicted both total cases and geographic spread, whereas the lower-end calibration best reproduced the observed outbreak size and spatial confinement. This supports transmission in this outbreak operating toward the lower end of measles' transmissibility range within a spatially clustered, partially immune population. We did not, however, systematically vary the within-school and household per-contact probabilities that set the baseline reproduction number, and counterfactual burden estimates should therefore be interpreted as conditional on this calibrated transmissibility. The realized reproductive number in any given simulation also varies stochastically with contact network structure, household composition, and local vaccination coverage.

The household transmission probability ($p_{\text{hh}}=0.25$ per susceptible contact per day) produces an expected household secondary attack rate of approximately 90% per susceptible co-resident over the 8-day infectious period, consistent with reported measles secondary attack rates in close-contact household settings. This figure assumes independent daily exposure across the infectious period; in small households, where repeated exposure to the same infectious individual is correlated, this independence assumption is an approximation.

**Vaccination Model**

Vaccination was implemented as an all-or-nothing model with efficacy $V_{E}=0.97$ (5). Vaccination decisions were made at the household level to capture the clustering of unvaccinated children within families. For each household, a vaccination probability was computed as the enrollment-weighted average of reported vaccination coverage rates across all schools attended by children in that household. A single Bernoulli draw was then made for the household, and all school-age children received the same vaccination status, ensuring that siblings attending different schools are jointly vaccinated or jointly unvaccinated. Students without household assignments received independent draws at their school's reported coverage rate.

Vaccinated individuals entered state V. Among vaccinated individuals, a fraction $1-V_{E}$ were assigned primary vaccine failure, rendering them susceptible to infection; upon infection, these individuals transmitted at reduced infectiousness (reduction factor $\eta$). Non-student household members were assigned vaccination status correlated with the household's student vaccination outcome: if any child in the household was unvaccinated, non-school-age siblings were unvaccinated with probability $p_{\text{sib}}=0.8$ and adults with probability $p_{\text{adult}}=0.5$; if all children were vaccinated, non-student members were vaccinated independently at a baseline adult coverage rate of 0.95.

The choice of an all-or-nothing model over a leaky formulation was made in consultation with CDC measles modelers, who have adopted a similar approach in their Measles Outbreak Simulator. For school-level surveillance data reporting only aggregate coverage, either formulation can reproduce the observed population-level immunity, but the two differ in how breakthrough infections accumulate under repeated exposure. Under a leaky model, each vaccinated individual retains partial per-contact susceptibility, so cumulative infection probability compounds across the intense, repeated exposures characteristic of household transmission, tending to overestimate breakthrough infections in high-contact settings. The all-or-nothing model instead designates a fully protected fraction, avoiding this overaccumulation and yielding breakthrough estimates more consistent with the small number observed in this outbreak.

**Public Health Interventions**

The model incorporated two categories of public health interventions: case isolation and contact quarantine. Case isolation was triggered upon transition to the rash stage, with detection delay depending on the case's surveillance context. The first detected case in each school (the school index case) was isolated after a delay of $d_{\text{index}}=4$ days following rash onset, reflecting the time required for clinical recognition in the absence of established school-level surveillance. All subsequent cases in the same school were isolated at the same default delay ($d_{\text{secondary}}=4$ days from rash onset), reflecting the structured case identification protocol maintained throughout the response. Isolated individuals were removed from within-school and between-school transmission for the duration of their isolation period ($T_{\text{iso}}=21$ days) but remained present in their household and continued to contribute to household transmission, reflecting the practical reality that home isolation concentrates infectious individuals with susceptible co-residents.

To evaluate the potential impact of more rapid case detection, we performed a scenario analysis in which the isolation delay was reduced to $d=2$ days for all cases. This value was motivated by epidemiological monitoring of the 2025 South Carolina outbreak, which documented that the interval between symptom onset and isolation gradually improved from approximately 4 days during the early phase of the response to approximately 2 days during the later phase as surveillance capacity and clinical awareness increased. The 2-day scenario, therefore, represents a counterfactual in which this improved detection speed was achieved uniformly from outbreak onset, allowing quantification of the additional cases that could have been prevented by earlier establishment of rapid case identification.

Contact quarantine was applied to unvaccinated contacts of newly-isolated cases, identified from the 7-day rolling contact history maintained per school. Vaccinated contacts were not quarantined, consistent with standard public health practice in which vaccination status exempts contacts from mandatory quarantine. Each eligible unvaccinated contact was quarantined independently with probability $\kappa=0.80$, reflecting the possibility that some contacts may be missed by public health authorities. Contacts not successfully quarantined remained in their current disease state and continued to participate in school-based transmission. Quarantined individuals in the prodromal stage were handled as isolated rather than quarantined, consistent with the operational reality that a symptomatic contact identified through tracing is managed as a confirmed or probable case rather than an asymptomatic contact. Quarantined susceptible individuals transitioned to state QS and quarantined latent individuals to state QE. Both states were fully removed from school-based transmission but continued to contribute to household transmission during the quarantine period. Quarantine lasted $T_{Q}=21$ days, after which uninfected individuals returned to their pre-quarantine state. Quarantined individuals who developed clinical symptoms during quarantine were transitioned to isolation, ensuring that infected contacts were eventually detected and removed from school transmission regardless of their initial quarantine status.

To evaluate sensitivity of outbreak outcomes to contact tracing capacity, we assessed quarantine efficacy values of $\kappa\in\{0.25,0.50,0.80,0.90\}$, spanning scenarios from severely limited contact tracing to near-complete identification of contacts. The value $\kappa=0.80$ was used as the default based on observed response capacity; lower values represent resource-constrained or delayed tracing, while $\kappa=0.90$ represents an optimistic upper bound on field performance.

**Mid-Outbreak Initialization and Forecasting**

The model was developed and deployed in successive versions as the outbreak progressed and additional structure was incorporated. An initial school-level outbreak risk tool was deployed at week 9, an inter-school transmission model at week 17, and the full household-integrated model at week 34, each shared with response leadership as surveillance data accumulated. Week numbering is relative to the first confirmed case. Each version was initialized from the cumulative surveillance state at the time of deployment, and forecasts were updated as new case and quarantine data became available. The retrospective validation reported in the main text uses the final household-integrated model initialized from outbreak onset, enabling consistent evaluation of model performance and intervention impact across the full outbreak.

To support real-time outbreak forecasting, the model includes a mid-outbreak initialization mode that seeds simulations from observed surveillance data rather than from a single index case. The required inputs are: (1) a list of schools that have reported confirmed exposures; (2) the cumulative number of confirmed cases to date; (3) the number of contacts currently in quarantine; (4) an estimate of the fraction of confirmed cases that remain actively infectious, derived from the distribution of illness onset dates relative to the initialization date; and (5) an estimated household secondary attack rate used to seed household members in affected households at initialization.

Confirmed cases were distributed across exposed schools in proportion to each school's unvaccinated susceptible pool size, reflecting the assumption that transmission risk scales with local susceptibility. The total case count was partitioned into recovered and active individuals using the provided active fraction. Active cases were further distributed across disease stages using fixed proportions informed by stage durations: 20% latent (E), 35% prodromal (P), and 45% rash-infectious ($R_{a}$). Active individuals were initialized at a uniformly sampled time point within their respective disease stage, capturing the range of progression states present in an ongoing outbreak. Quarantine contacts were initialized as partially elapsed, with time-in-quarantine drawn uniformly from 0 to 7 days, reflecting that contacts identified by an active response are at various stages of their quarantine period.

To ensure the correct application of isolation delays in mid-outbreak scenarios, a recovered individual from each exposed school was designated as the school index case. This designation signals to the model that school-level surveillance is already established, so all active cases receive the secondary isolation delay rather than the longer index case delay.

Non-student household members of students in exposed schools were seeded at initialization by applying the provided household secondary attack rate to each susceptible household contact independently, with the resulting infected household members distributed across active and recovered stages using the same proportions applied to school agents. This step ensures that the household transmission pool at simulation start reflects cumulative household exposure during the period preceding initialization rather than an unexposed household population.

The model was used prospectively to forecast total case counts, outbreak duration, and number of schools affected. Each scenario was run for 1,000 simulation replicates over a 365-day horizon, sufficient to capture complete outbreak extinction under all parameterizations examined, with outputs summarized as medians and prediction intervals across replicates.

**References**

1.	SC Immunization Data | South Carolina Department of Public Health. Available at: https://dph.sc.gov/public/vaccinations/sc-immunization-data [Accessed 21 May 2026].

2.	J. Rineer, *et al.*, A national synthetic populations dataset for the United States. *Sci Data* **12**, 144 (2025).

3.	Analyzing Midwest Social Contact Patterns - Modeling and Analytic Activities - School of Public Health - University of Minnesota. *School of Public Health*. Available at: https://www.sph.umn.edu/research/centers/midwest-analytics-and-disease-modeling/tools/analyzing-midwest-social-contact-patterns/ [Accessed 21 May 2026].

4.	F. M. Guerra, *et al.*, The basic reproduction number (R0) of measles: a systematic review. *The Lancet Infectious Diseases* **17**, e420–e428 (2017).

5.	CDC, Measles Vaccine Recommendations. *Measles (Rubeola)* (2026). Available at: https://www.cdc.gov/measles/hcp/vaccine-considerations/index.html [Accessed 21 May 2026].
